# A Multivariate Forecasting Model for the COVID-19 Hospital Census Based on Local Infection Incidence

**DOI:** 10.1101/2021.02.18.21251243

**Authors:** Hieu M. Nguyen, Philip Turk, Andrew McWilliams

## Abstract

COVID-19 has been one of the most serious global health crises in world history. During the pandemic, healthcare systems require accurate forecasts for key resources to guide preparation for patient surges. Forecasting the COVID-19 hospital census is among the most important planning decisions to ensure adequate staffing, number of beds, intensive care units, and vital equipment. In the literature, only a few papers have approached this problem from a multivariate time-series approach incorporating leading indicators for the hospital census. In this paper, we propose to use a leading indicator, the local COVID-19 infection incidence, together with the COVID-19 hospital census in a multivariate framework using a Vector Error Correction model (VECM) and aim to forecast the COVID-19 hospital census for the next 7 days. The model is also applied to produce scenario-based 60-day forecasts based on different trajectories of the pandemic. With several hypothesis tests and model diagnostics, we confirm that the two time-series have a cointegration relationship, which serves as an important predictor. Other diagnostics demonstrate the goodness-of-fit of the model. Using time-series cross-validation, we can estimate the out-of-sample Mean Absolute Percentage Error (MAPE). The model has a median MAPE of 5.9%, which is lower than the 6.6% median MAPE from a univariate Autoregressive Integrated Moving Average model. In the application of scenario-based long-term forecasting, future census exhibits concave trajectories with peaks lagging 2-3 weeks later than the peak infection incidence. Our findings show that the local COVID-19 infection incidence can be successfully incorporated into a VECM with the COVID-19 hospital census to improve upon existing forecast models, and to deliver accurate short-term forecasts and realistic scenario-based long-term trajectories to help healthcare systems leaders in their decision making.

**Author summary:** During the COVID-19 pandemic, healthcare systems need to have adequate resources to accommodate demand from COVID-19 cases. One of the most important metrics for planning is the COVID-19 hospital census. Only a few papers make use of leading indicators within multivariate time-series models for this problem. We incorporated a leading indicator, the local COVID-19 infection incidence, together with the COVID-19 hospital census in a multivariate framework called the Vector Error Correction model to make 7-day-ahead forecasts. This model is also applied to produce 60-day scenario forecasts based on different trajectories of the pandemic. We find that the two time-series have a stable long-run relationship. The model has a good fit to the data and good forecast performance in comparison with a more traditional model using the census data alone. When applied to different 60-day scenarios of the pandemic, the census forecasts show concave trajectories that peak 2-3 weeks later than the infection incidence. Our paper presents this new model for accurate short-term forecasts and realistic scenario-based long-term forecasts of the COVID-19 hospital census to help healthcare systems in their decision making. Our findings suggest using the local COVID-19 infection incidence data can improve and extend more traditional forecasting models.

## Introduction

SARS-Cov-2 is a novel member of the coronavirus family and infections in humans can result in corona virus disease of 2019, also known as COVID-19. The virus is transmitted primarily through droplets from coughing and sneezing and is highly infectious. Its basic reproduction rate is estimated to be in the low-to mid-2s based on different models [1], compared to 2 for SARS and 1.3 for 2009 H1N1 flu [2]. Moderate to severe disease typically manifests with acute hypoxemia, and can progress to acute respiratory distress syndrome, multi-organ dysfunction, and death. Furthermore, an estimated 25-30% of patients admitted to hospitals require intensive care admission [2]. In December 2019, the first cases were recorded in Wuhan, China with subsequent spread across the world. In early 2020, the World Health Organization declared COVID-19 to be a global health emergency [3]. At the end of December 2020, SARS-Cov-2 had resulted in over 82 million documented cases and nearly 2 million deaths [4].

Our work is motivated by the need from hospital leaders to have timely and accurate forecasts to guide planning for surges in hospital demands due to the pandemic. Adequate preparation can help prevent or mitigate strains on hospital resources COVID-19 that result when hospitals exceed their historical capacity. Thus, an essential tool is a model that provides short and long-range forecasting of the number of COVID-19 positive patients who will be admitted to a hospital or health system. This COVID-19 hospital census plays a central role in planning decisions that frequently require considerable lead-time, such as increasing staff, creating physical beds and rooms, and procuring vital equipment (e.g., ventilators and personal protective equipment).

Prior research has demonstrated the utility of forecasting hospital demands (e.g., hospital admissions, intensive care unit census, and hospital overall census) using univariate time-series models such as Autoregressive Integrated Moving Average (ARIMA), Seasonal Autoregressive Integrated Moving Average (SARIMA), and exponential smoothing [5–7]. Another approach is to use ensemble-based modeling. For example, a hybrid of a SARIMA model and a nonlinear autoregression artificial neural network model has been used to forecast hospital admissions [8]. In another example, two separate models, a time-series model for the hospital admission and a patient-level logistic regression model for the hospital discharge, were combined to predict the hospital census [9]. While these examples demonstrate the powerful potential of ensemble modeling, neither incorporate factors inherent to the behavior of the pandemic, which may serve as important leading indicators of hospital census, especially at times when infection rates become increasingly dynamic (e.g., on the approach or decent of peak infection prevalence). To incorporate pandemic indicators into modeling requires recognition that such indicators are typically nonstationary. Consequently, while a stationary multivariate time-series model, called Vector Autoregression (VAR), has been successfully employed to forecast emergency department patient census by including other hospital resource indicators [10], it cannot be used in this situation. Rather, our problem will require nonstationary multivariate time-series models like the Vector Error Correction model (VECM).

Recently, VECM has been used to forecast the demand for intensive care units during COVID-19 pandemic, by including the hospital admission as a leading indicator [11]. Although hospital admission is a natural choice of a leading indicator, it has a short period of lead time (i.e., hours to days) and thus, limited predictive power. A more powerful indicator for planning purposes would lead by days to weeks. We have previously used VECM to forecast COVID-19 hospital census using leading indicators from Google relative search volumes for COVID-19 testing related terms combined with the number of people flagged as having possible COVID-19 when using an internet-based virtual health screening bot [12]. However, these COVID-19 indicators, which are based on symptoms have limitations. For example, the symptoms of COVID-19 cannot be easily separated from other common conditions, such as the seasonal flu and search patterns may change due to other external factors over time.

A potentially more reliable and stable leading indicator for the COVID-19 hospital demand is the local COVID-19 infection incidence, which is the daily number of people officially confirmed to be COVID-19 positive in a defined area, in this case the target market of a hospital or a healthcare system. Because hospitalization typically lags by days to more than a week the onset of initial symptoms or exposure that may provoke a person to be tested, we hypothesized that at a local population level, infection incidence rates may serve as a leading indicator for predicting hospital census. In this paper, our main objective is to develop a multivariate time-series model that incorporates local COVID-19 infection incidence and COVID-19 hospital census, and explore whether the model has satisfactory 7-day-ahead forecast performance on COVID-19 hospital census.

## Materials and methods

### Time-series data

Atrium Health is a large, integrated healthcare system operating in North Carolina, South Carolina, and Georgia. In this paper, the COVID-19 hospital census (Census) is the daily aggregate number of beds occupied by COVID-19 patients, across the subset of 11 Atrium Health hospitals in the greater Charlotte metropolitan area of North Carolina, plus a virtual hospital (Atrium Health Hospital at Home). The local COVID-19 infection incidence (Incidence) is the aggregate daily count of new COVID-19 positive cases from 11 local counties belonging to CRI region (Cities Readiness Initiative), as designated by the North Carolina Department of Health and Human Services. The CRI region roughly approximates the market catchment area of these hospitals.

Using STL time-series decomposition [13], we observed that the 2 time-series had multiplicative weekly seasonality. We transformed both time-series to achieve periodic seasonality and linearize their relationship. The usual log transformation was applied to Incidence. For forecasting purposes, the health system had previously decided to place an upper bound of 1000 COVID-19 patients on the hospital time series range, so we applied a constrained log transformation (Equation (1)) so that the back-transformed Census (Equation (2)) forecasts would satisfy the constraint. The forecast model described in the following sections was developed for these transformed time-series. Fig 1 shows a plot of transformed Census and Incidence on a standardized scale for the period from May 15, 2020 to December 5, 2020. To affirm the association between the 2 transformed time-series, we computed the Pearson cross-correlations between (transformed) Census and values of (transformed) Incidence at lags 0, −1, …., −21.

**Fig 1.**
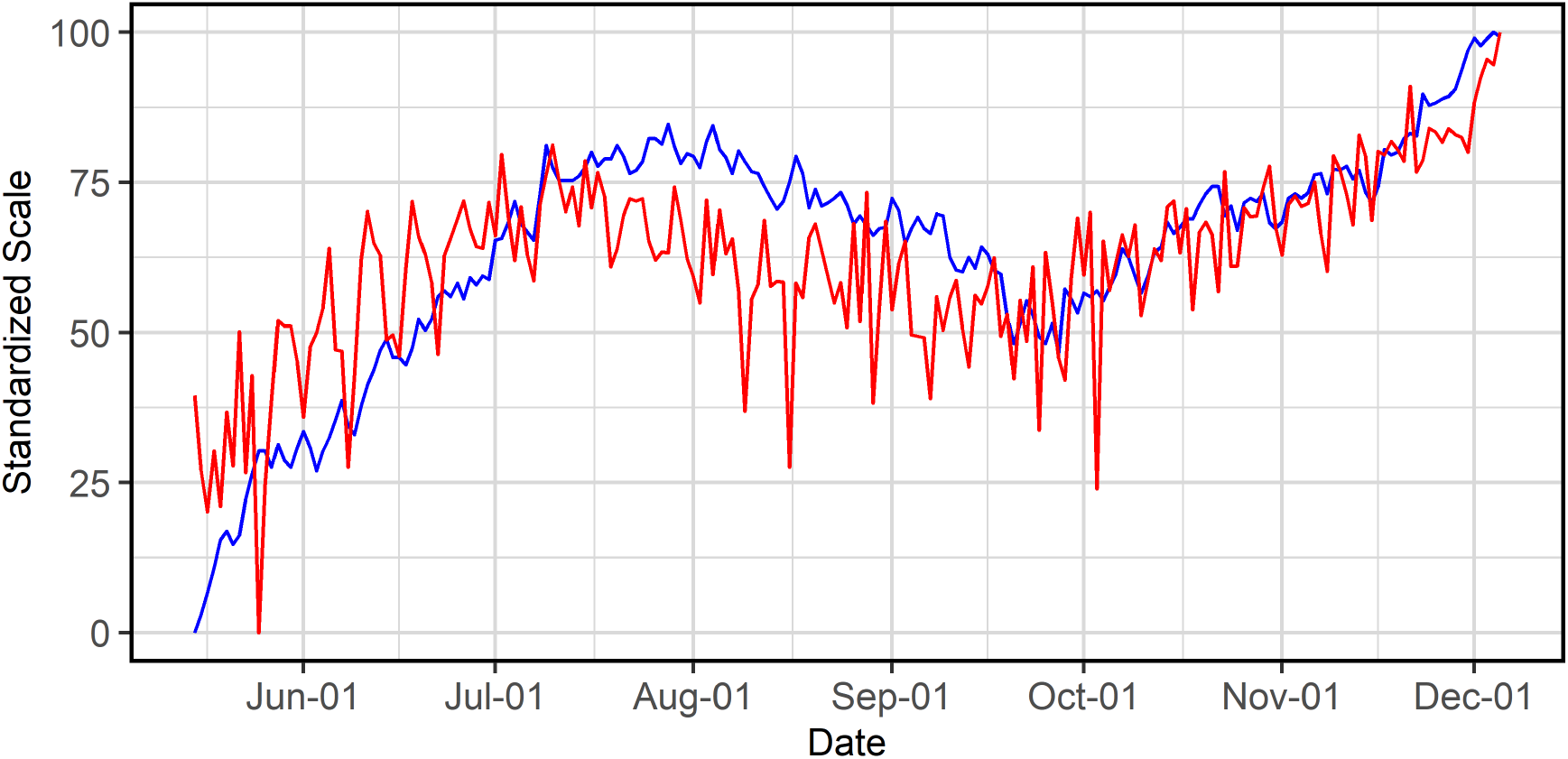
Scaled time-series for Census and Incidence for the period May 15, 2020 - December 5, 2020. Transformed Census (blue) and Incidence (red) are linearly standardized to the 0-100 scale.

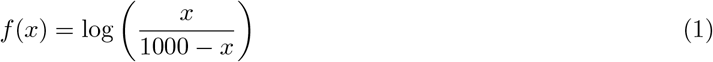

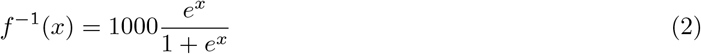

### Vector Error Correction model

A Vector Error Correction model is a vector autoregressive model used for nonstationary multivariate time-series and accounts for stable long-run relationships, i.e., cointegration, between the time series. A *k* x 1 time-series vector ***y***_*t*_ is said to be cointegrated if there is at least one non-zero *k* x 1 vector ***β***_*i*_ such that the linear combination 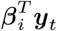 is trend-stationary. If *r* such linearly independent vectors ***β***_*i*_(*i* = 1, …, *r*) exist, we say ***y***_*t*_ is cointegrated with cointegration rank *r* [14].

Following [14], we first describe the VAR representation of order *p* of the VECM:

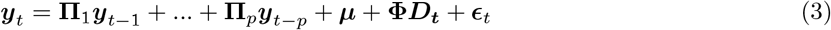

for time *t* = 1, …, *T*, where **Π**_*i*_ (for *i* = 1, …, *p*) are *k* x *k* coefficient matrices of the lagged series at lag *i*, ***µ*** is a *k* x 1 vector of constants, ***D***_*t*_ is a 6 x 1 vector of weekly seasonal indicators, **Φ** is a *k* x 6 coefficient matrix for seasonal indicators, and ***E***_*t*_ is a *k* x 1 vector of random errors.

The VECM specification can be formulated as an algebraic rearrangement of Equation (3):

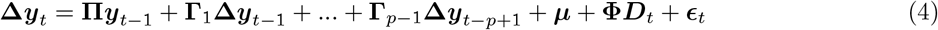

where **Δ*y***_*t*_ is a *k* x 1 vector of the differenced series ***y***_*t*_ *−* ***y***_*t−*1_, **Π** = *−*(***I*** *−* **Π**_1_ *−* … *−* **Π**_*p*_), and **Π**_*i*_ = *−*(**Π**_*i*+1_ + … + **Π**_*p*_) (for *i* = 1, …, *p −* 1).

The model has the following assumptions:

Assumption 1: The components of ***y***_*t*_ are at most *I*(1), i.e., integrated of order 1.

Assumption 2: 0 *≤ r* = *rank*(**Π**) *≤ k*

Assumption 3: ***E***_*t*_ are identically and independently distributed *N* (0, **Σ**) random vectors with covariance matrix **Σ**.

We now discuss the implications of the assumptions. For assumption 2, if *r* = *k*, then it can be shown that the VECM becomes a standard VAR model. If *r* = 0, **Π** is the zero matrix and there is no cointegration relationship between the series. The VECM then becomes a VAR model for first-differenced time-series. If 0 *< r < k*, then **Π** can be factored into **Π** = ***αβ***^*T*^, where ***α*** and ***β*** are both *k* x *r* matrices. From assumption 1, the differenced series **Δ*y***_*t*_, and its lags **Δ*y***_*t−*1_, …, **Δ*y***_*t−p*+1_, are stationary, and with Equation (4), it follows that **Π*y***_*t−*1_ = ***αβ***^*T*^ ***y***_*t−*1_ is (trend-)stationary, depending on the specification of the deterministic components. Therefore, ***β***^*T*^ ***y***_*t* 1_, also called the error correction term, is (trend-)stationary. The *r* linearly independent columns of ***β*** are the cointegrating vectors and the rank *r* is equal to the cointegration rank of ***y***_*t*_. In our case, since *k* = 2, we can have at most 1 cointegration relationship.

### Estimation and inference

The VECM was specified and fitted with the following steps:

First, to choose the order *p* of the VAR representation, we fitted a VAR model to the data and made the decision based on the Akaike Information Criteria (AIC) [15].

Second, we determined the number of cointegration relationships (*r* = 0 or *r* = 1) using the Johansen trace test [16].

Third, we needed to decide where to place the constant ***µ*** in the model. One option was to leave ***µ*** as shown previously to account for linear trend in the data. Another option was to restrict ***µ*** = ***αρ***. The constant would be absorbed into the cointegration relationship as an intercept and the data would not exhibit linear trend. The VECM specification then became:

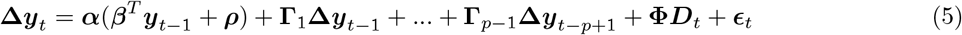

We made our decision about whether to restrict ***µ*** based on a likelihood ratio test for linear trend described in [17,18].

Fourth, we used maximum likelihood estimation to fit the model, reported parameter estimates, the corresponding *T* -tests, and the omnibus *F* -tests with the significance level of 0.05, following [17].

Finally, we computed the 7-day-ahead forecasts and the 80% forecast intervals. Once the forecasts of the transformed Census were made with the VECM, they were back-transformed to the original scale of Census with Equation (2). We created 80% forecast intervals for the transformed Census using a bootstrap procedure [19]. Then the lower and upper bound of the forecast intervals were also back-transformed with Equation (2).

The model was fitted to the data between May 15, 2020 and December 5, 2020. All the data analysis was done using R statistical software, version 4.0.3. The implementation of the VECM was done with the **tsDyn, vars** and **urca** R packages. Since there were no packages to make bootstrapped forecast intervals for the VECM, we coded our own implementation.

### Model diagnostics

We examined the omnibus *F* -tests to look for signs of lack of fit and also performed the multivariate Portmanteau test for the existence of serial correlation in the errors. Autocorrelation function and cross-correlation function plots were also generated for visual inspection. We performed the univariate and multivariate Jarque-Bera normality test on the errors [20] and also checked whether the cointegration relationship was stable, i.e., stationary, using the Augmented Dickey-Fuller (ADF) test [21] and the Kwiatkowski-Phillips-Schmidt-Shin (KPSS) test [22]. Finally, we checked the stability of the estimated VAR representation. To do so, we looked at the companion matrix of the VAR representation and checked whether the maximum eigenvalue modulus was strictly smaller than 1, which, if true, would imply the stability of the VAR representation [23]. We also generated a trace plot of the maximum eigenvalue modulus, where the model was repeatedly fitted on a daily rolling basis, to check for the consistency of this value over time.

### Forecast performance

We used Mean Absolute Percentage Error (MAPE) to evaluate the 7-day-ahead forecasts of Census:

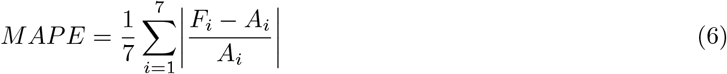

where *F*_*i*_ is the forecast value and *A*_*i*_ is the actual value.

In order to approximate the sampling distribution of MAPE, we performed time-series cross-validation. From June 16, 2020 to November 28, 2020, for each day, we iteratively fitted the model, made 7-day-ahead forecasts, and computed the MAPE. Eventually, we obtained 166 values of MAPE, plotted the distribution, and computed the median as well as the 95_th_ percentile.

### Scenario-based long-term forecasting

Leading up to and at the peak of infection prevalence, there can be high anxiety and uncertainty about how much more Incidence and, in particular, Census may increase. Furthermore, traditional univariate time-series models may give linear forecasts for Census that do not accurately represent pandemic behavior. However, cointegration allows for Census forecasts that leverage subtle, but critical, changes in Incidence (e.g., concavity). This suggests, if not necessitates, the forecasting of Census under different pandemic scenarios. In particular for resource planning, hospital leaders wish to understand the implications associated with a worst-case scenario.

For our healthcare system, besides routine 7-day-ahead Census forecasts, we also deployed our model for 60-day-ahead Census forecasts, considering 3 different scenarios of what could happen with Incidence, i.e., best-case, base-case, and worst-case. On January 9, 2021, we expected the winter surge to reach peak infection prevalence around February 5, 2021 based on an extension of an epidemiological model called Susceptible-Infected-Recovered [24]. While peak infection incidence typically leads peak infection prevalence, in the absence of definitively knowing either peak date, we took a conservative approach and linearly extrapolated Incidence with positive trend up to the expected pandemic peak. The severity of a scenario was controlled by a trend-dampening parameter [25]. After the peak, the descent path was initially symmetric to its accent then eventually became linear (Fig 2).

**Fig 2.**
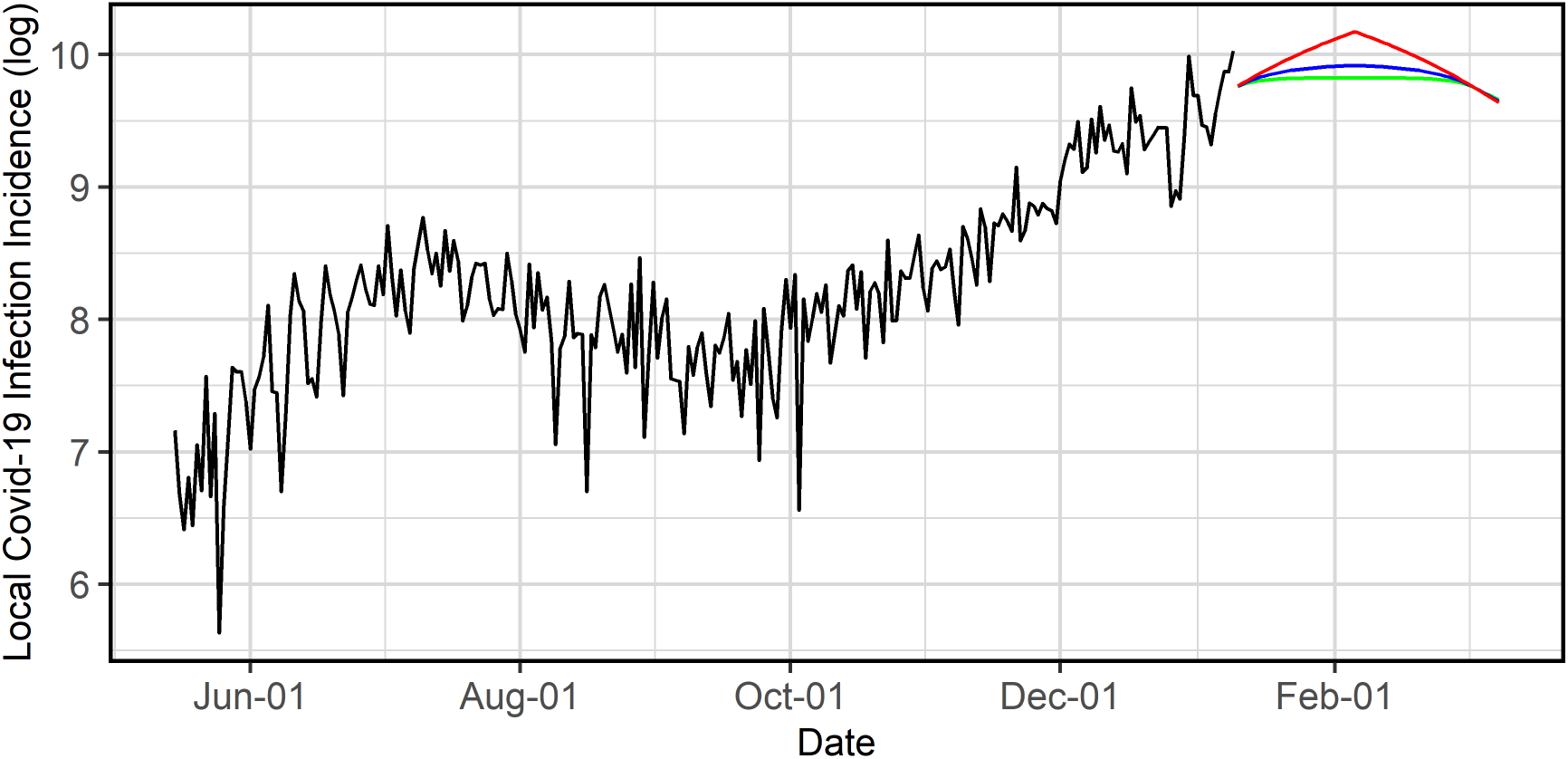
60-day projected local Covid-19 infection incidence on the log scale, as of January 9, 2021. Past values (black), worst-case scenario (red), base-case scenario (blue), best-case scenario (green).

Our model, refitted on January 9, 2021 with an increased capacity of 1250 patients, created forecasts iteratively forward for 60 days using the past Census forecasts together with projected Incidence under each scenario. To account for uncertainty in future Census and Incidence, we also simulated 1000 conditional sample paths of the two time-series under each scenario using the bootstrap procedure mentioned earlier and computed the 10^th^ and 90^th^ percentile at each horizon to obtain the 80% forecast intervals.

## Results

### Estimation and inference

Our model was specified as a VECM with 7 lags in its VAR representation (*p* = 7), 1 cointegration relationship (*r* = 1), and a restricted constant parameter so that the series would not have linear trend. The AIC scores of VAR models with different number of lags from 2 to 14 were inconclusive. However, we found that 7 lags were sufficient to account for all the correlation in the data, as evidenced by the autocorrelation function and cross-correlation function plots of the residuals (Fig 3). The Johansen trace test indicated that there was 1 cointegration relationship (p-value < 0.01). Finally, the likelihood ratio test for linear trend indicated that there was no linear trend in the data (p-value = 0.32). Furthermore, the restricted model had a lower AIC score than the unrestricted model (the AIC scores were −1519 and −1516, respectively).

**Fig 3.**
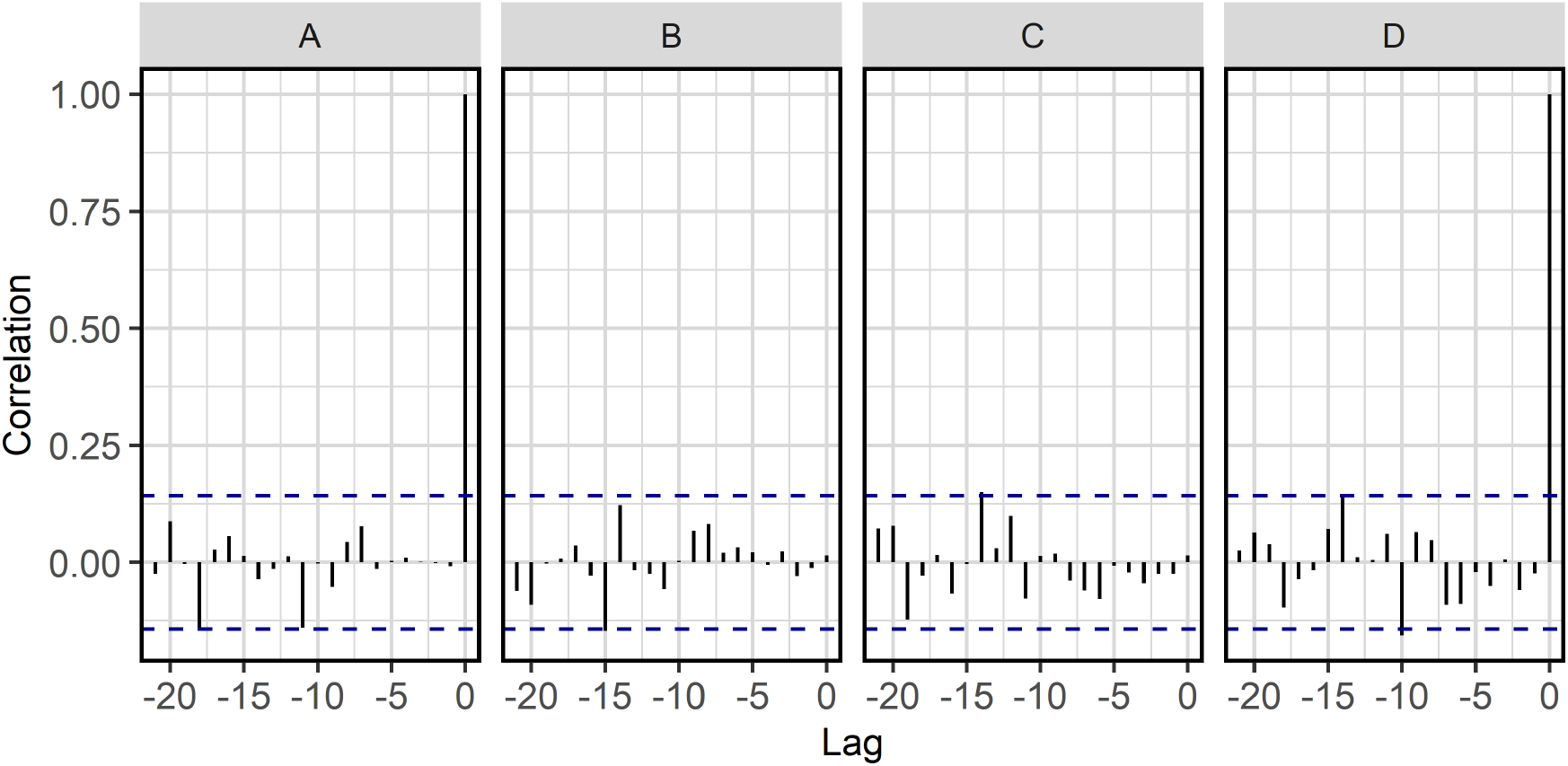
Autocorrelation functions and Cross-correlation functions of the residuals. (A) Census residuals, (B) Lagged Census residuals and Incidence residuals, (C) Census residuals and lagged Incidence residuals (D) Incidence residuals.

The outputs from the maximum likelihood estimation showed that the cointegration relationship, i.e., the error correction term, had a significant negative effect on Census change (p-value < 0.0001); no significant effect was observed for Incidence change (p-value = 0.2590) (Table 1). The long-run relationship was estimated as:

**Table 1.** Parameter estimates and *T* -tests.

| Predictor | $\Delta\text{Census}_t$ | | | $\Delta\text{Incidence}_t$ | | |
| --- | --- | --- | --- | --- | --- | --- |
| | Estimate | $T$-statistics | P-value | Estimate | $T$-statistics | P-value |
| Fri | -0.0213 | -1.1120 | 0.2676 | 0.0095 | 0.1024 | 0.9186 |
| Sat | 0.0083 | 0.3980 | 0.6911 | -0.1528 | -1.5176 | 0.1309 |
| Sun | 0.0030 | 0.1330 | 0.8943 | -0.3340 | -3.0744 | 0.0024 |
| Mon | 0.0585 | 2.6205 | 0.0095 | -0.1939 | -1.7950 | 0.0743 |
| Tue | 0.0291 | 1.3896 | 0.1664 | -0.1284 | -1.2655 | 0.2074 |
| Wed | -0.0037 | -0.1895 | 0.8499 | 0.0343 | 0.3672 | 0.7139 |
| $\text{ect}_{t-1}$ | -0.1265 | -5.6993 | 0.0000 | -0.1216 | -1.1323 | 0.2590 |
| $\Delta\text{Census}_{t-1}$ | -0.0489 | -0.7143 | 0.4760 | 0.5487 | 1.6555 | 0.0996 |
| $\Delta\text{Incidence}_{t-1}$ | -0.0665 | -2.8222 | 0.0053 | -0.9808 | -8.6067 | 0.0000 |
| $\Delta\text{Census}_{t-2}$ | -0.2220 | -3.2277 | 0.0015 | -0.0614 | -0.1844 | 0.8539 |
| $\Delta\text{Incidence}_{t-2}$ | -0.0532 | -2.0881 | 0.0382 | -0.6955 | -5.6431 | 0.0000 |
| $\Delta\text{Census}_{t-3}$ | -0.0700 | -0.9949 | 0.3211 | 0.0643 | 0.1890 | 0.8503 |
| $\Delta\text{Incidence}_{t-3}$ | -0.0472 | -1.9094 | 0.0578 | -0.6428 | -5.3755 | 0.0000 |
| $\Delta\text{Census}_{t-4}$ | -0.0785 | -1.1224 | 0.2632 | 0.9769 | 2.8871 | 0.0044 |
| $\Delta\text{Incidence}_{t-4}$ | -0.0567 | -2.4165 | 0.0167 | -0.5564 | -4.8999 | 0.0000 |
| $\Delta\text{Census}_{t-5}$ | -0.0499 | -0.7140 | 0.4762 | -0.0792 | -0.2341 | 0.8152 |
| $\Delta\text{Incidence}_{t-5}$ | -0.0465 | -2.1907 | 0.0298 | -0.4589 | -4.4634 | 0.0000 |
| $\Delta\text{Census}_{t-6}$ | 0.0077 | 0.1107 | 0.9120 | 0.4533 | 1.3404 | 0.1818 |
| $\Delta\text{Incidence}_{t-6}$ | -0.0373 | -2.4015 | 0.0174 | -0.2384 | -3.1739 | 0.0018 |

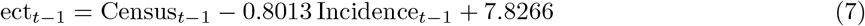

where ect_*t−*1_ was the (lagged) error correction term.

Table 1 also showed that past changes in Census and Incidence also had meaningful effects on current Census change. Past Census changes a had significant effect at lag 2 (p-value = 0.0015). Past Incidence changes had significant effects at lag 1 (p-value = 0.0053), lag 2 (p-value = 0.0382), lag 4 (p-value = 0.0167), lag 5 (p-value = 0.0298), and lag 6 (p-value = 0.0174).

There were some significant seasonal effects; that is, differences in both Census and Incidence changes among weekdays. Compared to Thursday, Census change was higher on Monday and Incidence change was lower on Sunday, with significant differences (p-value = 0.0095 and 0.0024, respectively).

### Model diagnostics

The omnibus *F* -tests were significant for both Census (p-value < 0.0001) and Incidence (p-value < 0.0001) components.

The Portmanteau test did not show sufficient evidence that the errors were autocorrelated (p-value = 0.1873). From the residual autocorrelation function and cross-correlation function plots, the correlations were within the 95% confidence band (Fig 3). The Jarque-Bera normality tests failed to reject the normality null hypothesis for the Census errors (p-value = 0.7100), but did for Incidence (p-value < 0.0001). Specifically, the Incidence residuals were moderately left-skewed. The Jarque-Bera multivariate test also rejected the multivariate normality null hypothesis (p-value < 0.0001).

The Augmented Dickey-Fuller test for stationarity of the error correction term rejected the unit root null hypothesis at 10% significance level but failed to reject the null hypothesis at 5% significance level. The KPSS test failed to reject the stationarity null hypothesis (p-value = 0.0985). Examination of the time plot of predicted ***β***^*T*^ ***y***_*t−*1_ showed no obvious departure from stationarity.

The companion matrix of the VAR representation had a maximum eigenvalue modulus of 0.97, strictly less than 1. Although this value was close to 1, the trace plot showed that this value had been steadily below 1 across time, when the model was fitted repeatedly in a daily rolling basis from June 16 to November 28 (Fig 4).

**Fig 4.**
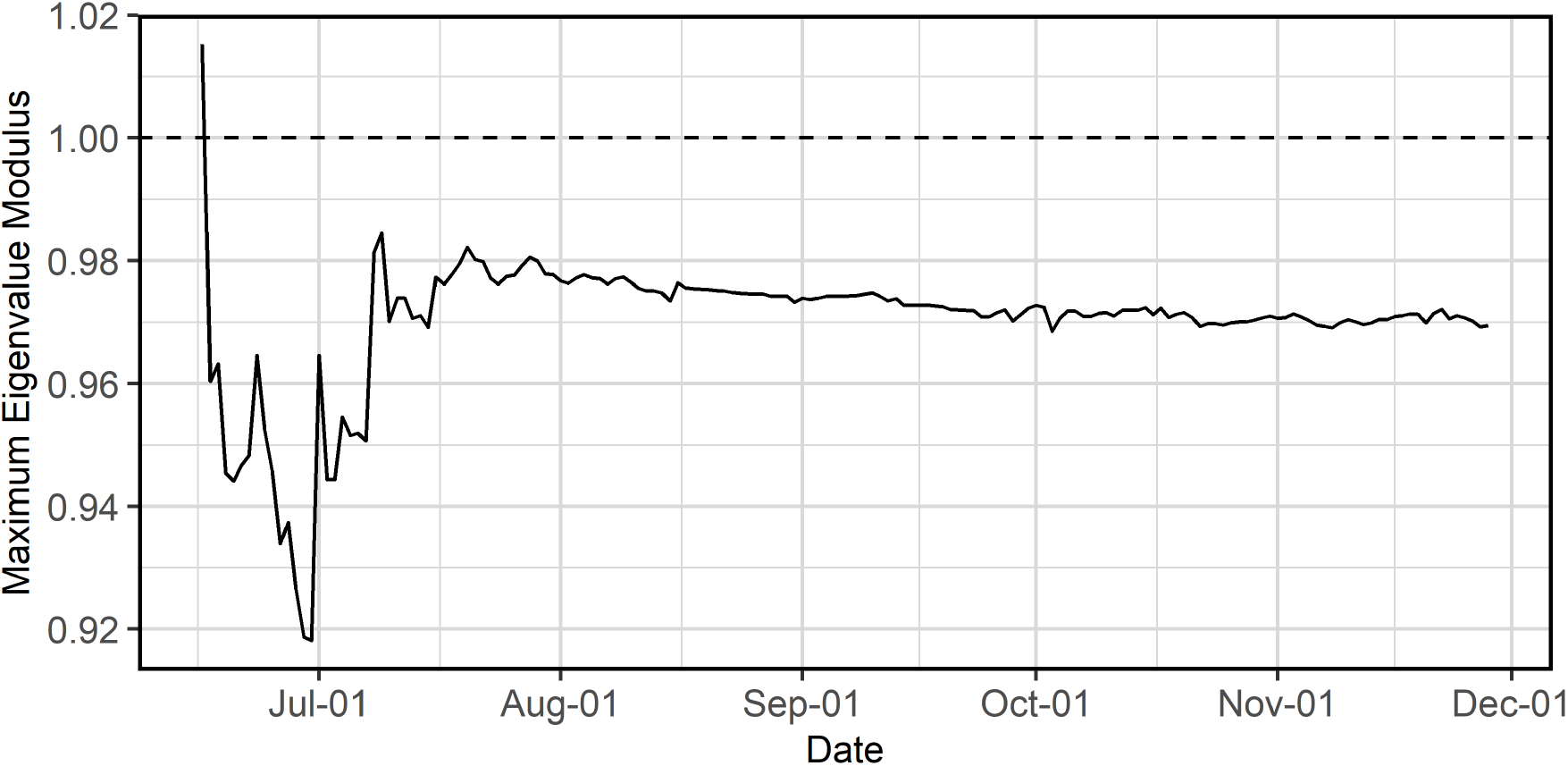
Trace plot of the maximum eigenvalue modulus for the period June 16, 2020 - November 28, 2020.

### Forecast performance

We obtained the approximate sampling distribution of the out-of-sample MAPE from time-series cross-validation (Fig 5). The typical value (median) of MAPE was 5.9% and the 95^th^ percentile of MAPE was 13.4%. For the sake of comparison, the corresponding values from an ARIMA model using the COVID-19 hospital census only were 6.6% and 14.3%. Additionally, after fitting the May 15 - December 5 data, we forecasted the Census out to 7 days. Subsequently, the actual values were accurately forecasted with a MAPE of 1.9% and were all within the 80% bootstrapped forecast intervals (Fig 6).

**Fig 5.**
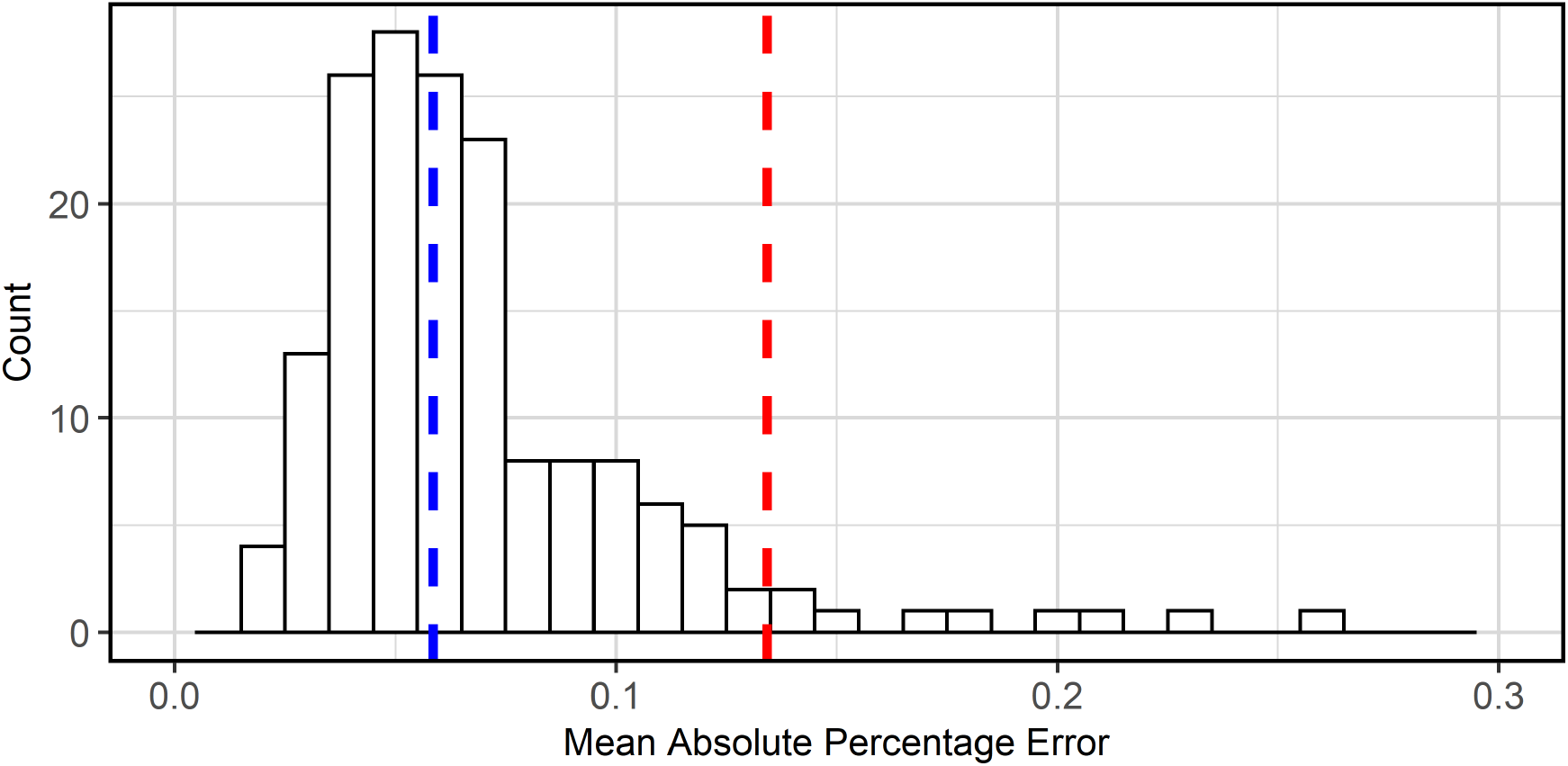
Distribution of the 7-day-ahead Mean Absolute Percentage Error from time-series cross-validation for the period June 16, 2020 - November 28, 2020. Median (blue), 95^th^ percentile (red).

**Fig 6.**
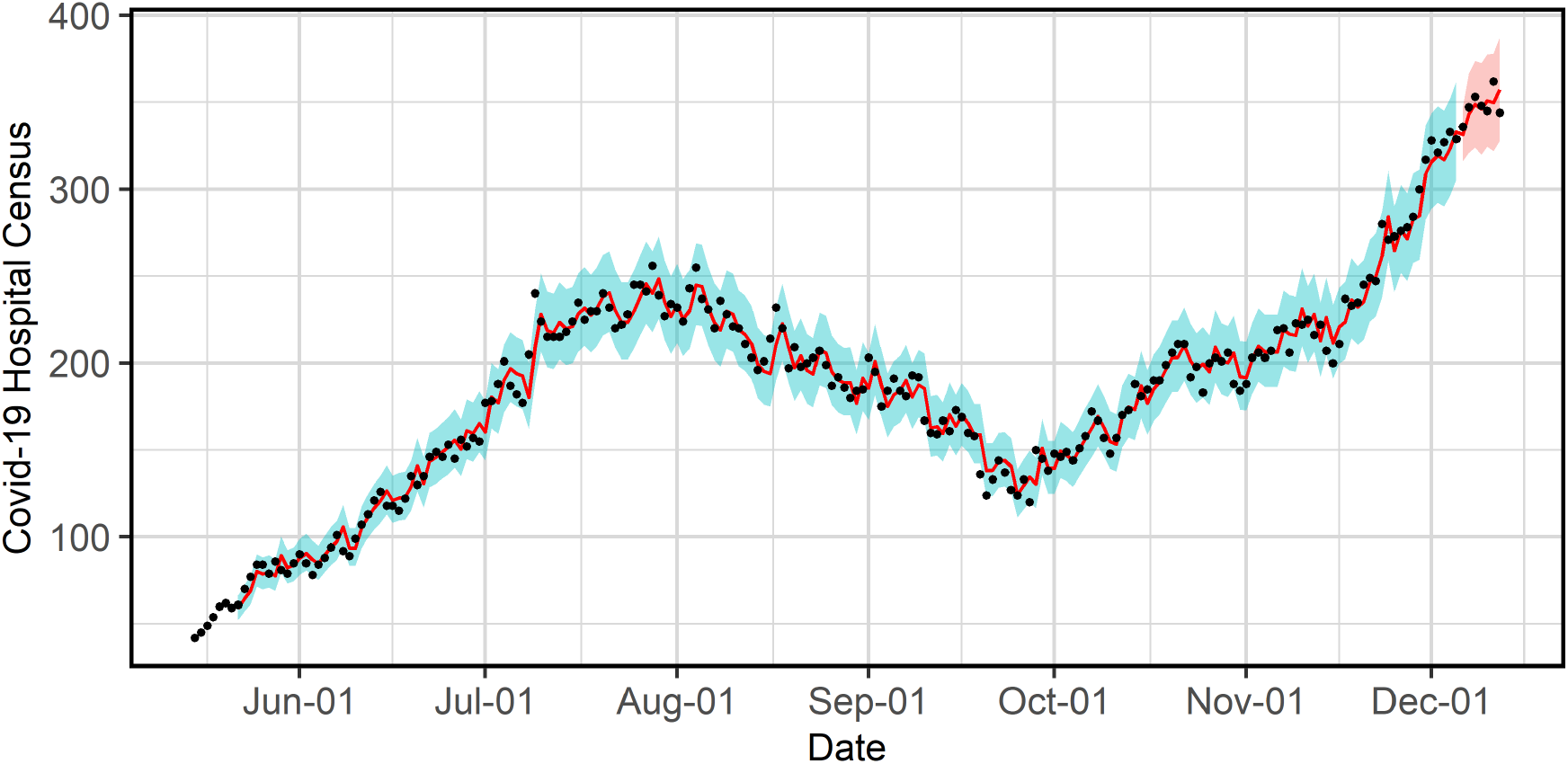
One-step-ahead in-sample and 7-day-ahead out-of-sample predictions. True values (black), in-sample and out-of-sample predictions (red line), 95% confidence intervals (blue band), 80% forecast intervals (red band). The model is fitted on data from May 15, 2020 to December 5, 2020.

### Scenario-based long-term forecasting

In all scenarios, due to cointegration, Census followed corresponding concave trajectories with peaks occurring approximately 2-3 weeks later than Incidence depending on the scenario. In the worst-case scenario, Census was projected to peak on February 16, 2021 (11 days later than Incidence) with approximately 850 patients at the 80% forecast interval upper bound (Fig 7).

**Fig 7.**
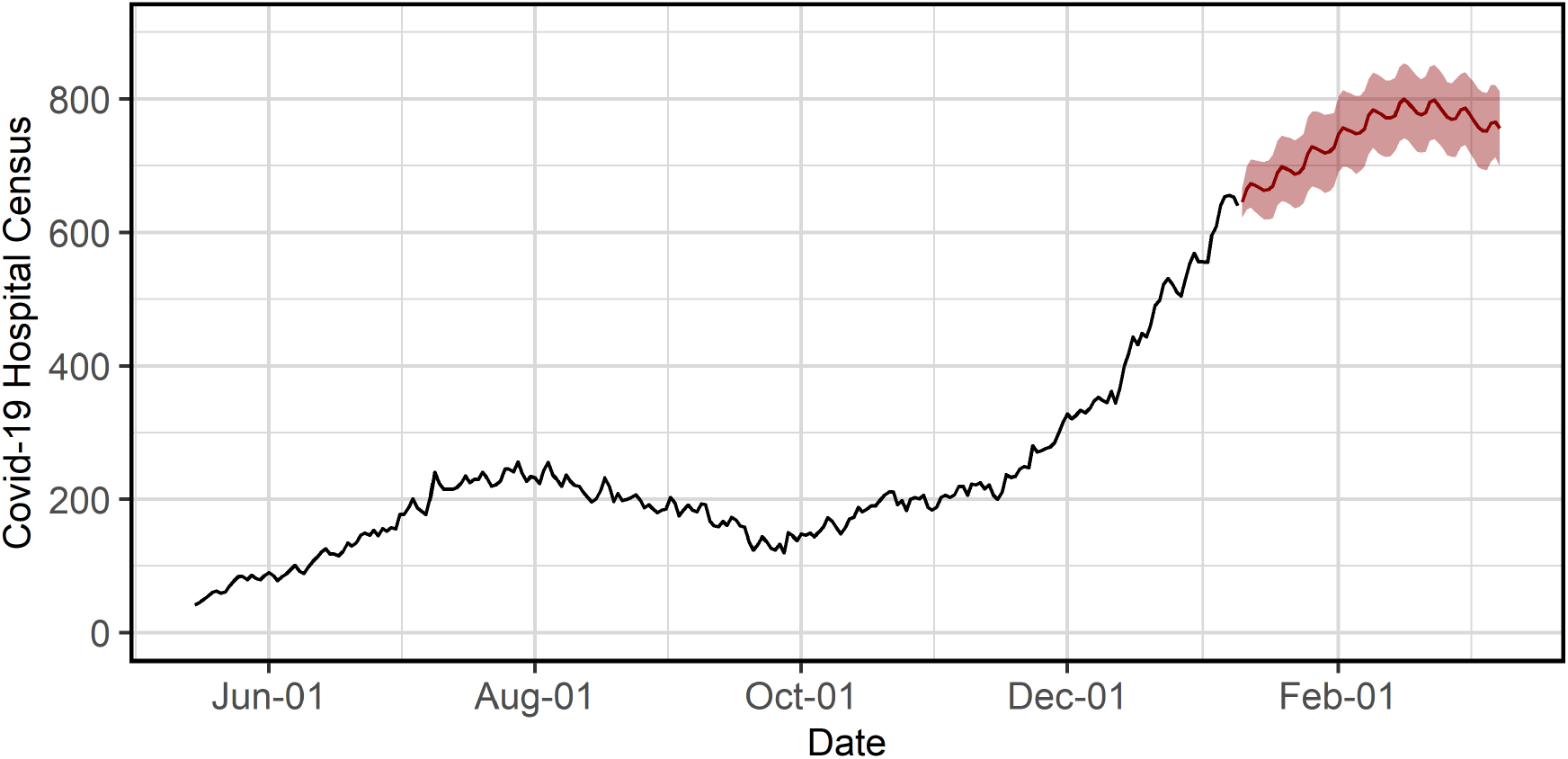
60-day Census forecasts in the worst-case scenario, as of January 9, 2021. Past values (black), forecasts (red line), 80% forecast intervals (red band).

## Discussion

Our VECM provides a very good fit to the data and outperforms models with no or other leading indicators. Significant omnibus *F* -tests showed that the model fit was better than that of a reduced VECM representation with no predictors, i.e., a bivariate random walk model. When we examined model diagnostics, there was no sign of any serious departure from model assumptions. From the Portmanteau test, the errors were not different from white noise. Although the normality assumption (for Incidence) was not met, the asymptotic properties of our estimation and hypothesis tests in the VECM would not be affected [26]. To address the possible effect of this violation on the forecast intervals, we implemented a bootstrap procedure for the forecast intervals. Both the ADF test and KPSS test showed reasonable evidence that the long-run relationship was stable. With the maximum eigenvalue modulus of the VAR representation consistently below 1 across time, the model itself was quite stable. In terms of forecast performance, the VECM yielded smaller MAPE, in terms of the median and the 95^th^ percentile, when compared to an ARIMA model using the COVID-19 hospital census only. Our VECM also performed better than another VECM that uses two Internet-based leading indicators (median MAPE of 10.5%), albeit on time domains that were partially overlapping [12].

The long-run relationship plays a crucial role in the model. Our model results show how future Census responds to perturbations in the long-run cointegration relationship in the direction that would preserve the stability of the relationship. For instance, if Incidence increases significantly and drives the error correction term below 0, the next day Census will tend to increase so that the error correction term will move back towards 0. Compared to short-run relationships between Census change and past changes in Incidence and Census, the long-run relationship effect is strongly significant and is a major driver in the model.

We observe that local infection incidence led the hospital census by about 2 weeks. The cross-correlation between Incidence and Census had uniformly high positive correlation, between 0.7 and 0.8 at different lags, but the highest correlation was at lag 14. Clinically, we know that after someone is diagnosed with SARS-CoV-2, it can take several days before they get sick enough to be hospitalized. During the summer 2020 wave of the pandemic, Incidence peaked 18 days earlier, on July 10, than when Census peaked, on July 28. In the model, we also saw that past Incidence changes at multiple lags have statistically significant effects on Census. In total, the model results and our observations demonstrate the predictive power of Incidence.

Applying the model to scenario-based forecasting is an important method at a healthcare system for long-term forecasting when approaching an infection prevalence peak and helps determine the potential for resource capacity to be exceeded under a worst-case scenario. Our method has several advantages compared to others. First, with a scenario-based and epidemiologically informed approach, our VECM produces realistic non-linear long-range trajectories of Census. In contrast, an ARIMA model can have an upward linear trajectory even as we approach and arrive at the infection prevalence peak because it is agnostic to Incidence. Hence, the VECM fit with scenario-based Incidence will provide better accuracy since it is more reflective of pandemic behavior. Second, other scenario-based models, such as the COVID-19 Hospital Impact Model for Epidemics (CHIME) at University of Pennsylvania [27], ignore the delay between hospitalization admission and infection incidence and let the proportion between the 2 variables be a time-invariant scenario parameter. Our model acknowledges this important temporal dynamic and provides empirical estimation. Third, when our concern is a specific scenario, our approach is particularly useful at minimizing long-range forecast uncertainty, since the bootstrapped sample paths are constrained to fluctuate around the marginalized scenario-based Census projection. Without such a constraint, 60-day forecasts can typically have wide forecast intervals that are of no practical utility.

Although our model has been thoroughly developed, it is not free of limitations. First, it is possible that we may lose the stable long-run relationship at some point in the future, due to structural changes in the time-series brought on by evolving pandemic behavior. The VECM can be modified to account for some basic types of changes such as level shifts [28,29]. However, modeling more complex structural changes have received limited research. Second, it is relatively more difficult to fit a VECM. For univariate models such as ARIMA and exponential smoothing, well-developed R packages exist for automated model specification and estimation. With the VECM, more deliberate modeling decisions and careful checking of assumptions need to be made to fit a reliable model. Third, the inclusion of seasonal effects in our model requires that the seasonality is periodic. However, another healthcare system may find that their time-series data have non-periodic seasonality or multiple periodic seasonality. If seasonality is not important, we may resolve this by simply de-seasonalizing the series. Otherwise, it may be possible to account for this with more advanced parameterization of the seasonal effects.

Our work demonstrates the potential of utilizing local infection data in a multivariate time-series VECM. The construct presented here provides a framework for incorporating other leading indicators that may yield further increases in the predictive power. For instance, the VECM that uses Internet-based leading indicators

[12] could potentially be improved by including Incidence. It is also possible to incorporate other nested hospital-related time-series, such as the number of intensive care units, and the number of ventilators, into the VECM if there was a need to simultaneously forecast other resources. Additionally, a VECM could be a valuable candidate for a model-averaged ensemble. This can be particularly useful if the ensemble consists only of agnostic univariate time-series models.

In summary, we have presented a model that successfully tethers infection incidence with hospital census and achieves accurate forecasting of COVID-19 hospital census, providing critical insights to guide health system decision-making. Other health systems can leverage this model, using local epidemiological and census data, to help navigate the pandemic.

## Data Availability

The data and code used in the data analysis is publicly available at GitHub.

https://github.com/hmnguye/Incidence-Census-Model

## Author contributions

Conceptualization: Hieu Nguyen, Philip Turk

Formal analysis: Hieu Nguyen, Philip Turk

Supervision: Andrew McWilliams

Writing – original draft preparation: Hieu Nguyen

Writing – review & editing: Hieu Nguyen, Philip Turk, Andrew McWilliams

## Notes

### Competing Interest Statement

The authors have declared no competing interest.

### Funding Statement

Neither the authors nor their institution received external funding for the conduct of this study.

### Author Declarations

Protocol was submitted to the Atrium Health Institutional Review Board (IRB) prior to execution and the study was deemed exempt from IRB oversight. In compliance with HIPAA regulations, individual patient information were not disclosed, and all these data have been deidentified and reported as aggregates. Other data are openly available to the public. Full names / affiliations of all ethics committees / Institutional Review Boards: Jon Schwaiger, CIP, Administrative IRB Director, Research Facility Privacy Officer. Decision made, i.e. whether ethical approval was given or waived: Waived.

